# Do Perspectives Matter? Comparing Patient, Informant, and Clinician Subjective Cognitive Decline

**DOI:** 10.64898/2026.02.13.26346246

**Authors:** Camille Barrette, Mahsa Dadar, Cassandra Morrison

**Affiliations:** Department of Psychology, Carleton University, Ottawa, ON, K1S 5B6, Canada; Department of Psychiatry, McGill University, Montreal, Quebec, H3A 1A1, Canada; Douglas Mental Health University Institute, Montreal, Quebec, H4H 1R3, Canada

**Author notes:** Co-senior authors (Mahsa Dadar, Cassandra Morrison). **Corresponding author**: Cassandra Morrison, Department of Psychology, Carleton University, Ottawa, ON, K1S 5B6, Canada.

**Keywords:** Subjective Cognitive Decline, Informant reports, Clinician Reports, Cognitive Change, Cognitive Performance

## Abstract

**BACKGROUND:** Patient reports are the standard when examining subjective cognitive decline (SCD). Recent research suggests that informant and clinician reports may also be associated with cognition. This study examined differences between patient, informant, and clinician definitions of SCD and their relationship to cognition.

**METHODS:** Data from 4290 older adults (n=1690 normal controls, NC; n=840 mild cognitive impairment, MCI; n=1760 Alzheimer’s disease, AD) were examined from the National Alzheimer’s Coordinating Center. Linear models examined the relationships between SCD status using the three definitions and cognition at baseline and over time.

**RESULTS:** In NC, informant and clinician SCD were associated with worse cognition at baseline, with patient and clinician SCD associated with worse cognition over time. All definitions were associated with worse cognition at baseline and over time in MCI and AD.

**DISCUSSION:** Our findings suggest the importance of examining different SCD definitions, especially the inclusion of clinician SCD.

## 1. Background

With a growing percentage of the population expected to convert to dementia in the coming years [1], more research is focusing on identifying early detection markers to predict and prevent cognitive decline [2]. Of the practical and widely applicable early markers which has proven to be increasingly promising is subjective cognitive decline (SCD) [2–5], defined as the subjective experience of a decrease in cognitive function in the absence of objective decline [6]. That is, individuals with SCD perform within the normal range for their age group, despite feeling as though their cognition has declined [2,3,6,7]. Previous research has investigated the relationship between SCD and cognitive decline, with many studies reporting that compared to individuals without SCD (SCD-), older adults with SCD (SCD+) are more likely to progress to mild cognitive impairment (MCI) or dementia [2,8–11].

The relationship between cognitive performance and SCD has generally shown that endorsement of SCD by an older adult is related to worse cognitive performance [12–15] and increased rates of cognitive decline over time [15–18]. While some studies have reported a link between objective performance and SCD on measures of global cognition [14–16,19,20], memory [14,15,18,20–22], executive function [18,22,23], and language [17], others have found no association in these domains [16,17,20,21,24]. These conflicting results may be related to the heterogeneity of SCD research across studies, with many different techniques, models, populations, and definitions of SCD employed[2,17,25], limiting the generalizability of current findings [14,17,25]. Therefore, further research is needed to enable more definitive conclusions on the relationship between SCD and cognitive performance and decline.

While SCD is typically defined by self-experienced cognitive decline [2], confirmation of decline from an informant or observer by someone close to the patient [2,3] or a clinical professional [26] have emerged as additional markers that could improve the relationship between SCD and cognitive performance. Informant reported SCD refers to when an outside informant or observer corroborates the patient’s concerns about the declining state of their cognition [6,27]. Informant reported SCD has been shown to be associated with further decline in cognitive performance [28,29], especially when paired with a self report of SCD [9,16,21,25,27,28], when individuals are older [8,30], and when individuals have MCI [2,19,31,32]. Informant SCD has also been suggested as a stronger predictor of further objective decline compared to self-report, with one meta-analysis reporting that while both self and informant reports were associated with an elevated risk of transition, the association was stronger for informant reported complaints [9]. Informant report was also added as a key SCD *plus* feature, meaning that those with a positive informant SCD report are at a greater risk of conversion to dementia [3,17]. In addition to informant reports, self-reports of SCD can also be corroborated by a clinician [26]. However, to our knowledge, no prior research has examined the cross-sectional and longitudinal relationships between clinician SCD and objective cognitive performance.

The current study primarily aims to explore differences in cognitive decline profiles between SCD+ and SCD-participants across three definitions of SCD (patient, informant, and clinician), and across three diagnostic categories (normal cognition (NC), MCI, and Alzheimer’s disease (AD)). We also aim to explore if cognitive decline occurs in specific domains for different types of SCD and diagnoses. For patient reported SCD, research has shown that these reports tend to be most associated with language [17] and memory [21,22], whereas for informant reported SCD, reports are strongly linked with executive function [2,27] and memory [27] declines. Therefore, we will further explore the relationship between domain-specific cognitive decline with patient, informant, and clinician reports of SCD.

## 2. Methods

### 2.1 National Alzheimer’s Coordinating Center Participants

Data were obtained from the NACC database, which included the NACC Uniform Data Set 3 (UDS3). Baseline data from individuals aged 55-80 with diagnoses of cognitively normal (NC), mild cognitive impairment (MCI), and Alzheimer’s Disease (AD) were included in the study. Cognitive status was calculated using the NACCTMCI and NACCETPR variables (representing the primary diagnosis) as well as Clinical Dementia Rating-Sum of Boxes (CDR_SB) scores, ensuring consistency in cognitive categorization. Participants with CDR_SB of 0 and no neurological disorders based on NACCETPR variable were classified as “NC”, those with a CDR_SB between 0.5 and 4 were classified as “MCI,” and those with CDR_SB > 4.5 were classified as “AD”. Participants with other neurological disorders (Parkinson’s disease, Lewy body dementia, frontotemporal dementia, etc.) or inconsistent CDR scores and diagnostic labels (e.g., individuals with CDR_SB > 4.5 and no diagnosis identified based on NACCETPR) were excluded. Education was quantified as years of formal schooling completed as a continuous numeric variable. It is important to note that subjective concerns in the MCI and AD group are subsequently accompanied by an objective decline in cognition due to the criteria we established for our cognitive status groups. Therefore, the definitions in these groups do not fit under the established criteria for SCD, which assert that SCD occurs in the absence of objective decline [3]. Thus, SCD in these two groups of participants is more so defined solely by the cognitive complaints/concerns from patients, informants, and clinician.

### 2.2 Participant Categorization

Participants were further selected based on the availability of baseline patient, informant, or clinician reported SCD, and the availability of scores for the cognitive measures evaluated. Patient SCD was determined using the baseline question (DECSUB), “Does the subject report a decline in memory (relative to previously attained abilities)”? Informant SCD was determined using the baseline question (DECIN), “Does the co-participant report a decline in subject’s memory (relative to previously attained abilities)”? Finally, clinician SCD was determined using the baseline question (DECCLIN), “Clinician believes there is a meaningful decline in memory, non-memory cognitive abilities, behavior, ability to manage his/ her affairs, or there are motor/movement changes.” For each type of SCD, participants were categorized as having either a positive report of SCD (e.g. ‘SCD+’) or a negative report of SCD (e.g. (‘SCD-’). Demographic information, separated by each diagnostic group and type of SCD can be found in Tables 1–3.

**Table 1.** Demographic and variable characteristics for all SCD definitions for older adults in the NC group.

| NCs | Patient |  | Informant |  | Clinician |  |
| --- | --- | --- | --- | --- | --- | --- |
|  | SCD- | SCD+ | SCD- | SCD+ | SCD- | SCD+ |
| Total number | 1232 | 458 | 1517 | 173 | 1625 | 65 |
| Age | 68.8 ± 6.8 | 68.6 ± 6.0 | 68.8 ± 6.8 | 68.6 ± 6.0 | 68.8 ± 6.8 | 67.9 ± 6.0 |
| Female Sex | 808 (66%) | 302 (66%) | 1016 (67%) | 94 (54%) | 1069 (66%) | 41 (63%) |
| Education | 15.5 ± 3.3 | 15.6 ± 2.8 | 15.4 ± 3.3 | 15.8 ± 2.8 | 15.5 ± 3.3 | 15.5 ± 2.8 |
| CDR | 0.01 ± 0.3 | 0 ± 0.05 | 0.01 ± 0.3 | 0 ± 0 | 0.01 ± 0.3 | 0 ± 0 |
| Animal Fluency | 20.06 ± 5.77 | 19.96 ± 5.82 | 20.11 ± 5.80 | 19.38 ± 5.54 | **20.10 ± 5.76** | **18.33 ± 6.00** |
| Vegetable Fluency | 14.69 ± 4.33 | 14.84 ± 4.16 | **14.81 ± 4.26** | **14.02 ± 4.49** | 14.76 ± 4.30 | 13.86 ± 3.70 |
| Trail-Making A | 33.16 ± 15.19 | 32.96 ± 13.17 | 33.02 ± 14.72 | 33.85 ± 14.37 | 32.99 ± 14.64 | 36.03 ± 15.53 |
| Trail-Making B | 75.93 ± 26.27 | 75.79 ± 25.14 | 75.65 ± 25.98 | 77.98 ± 25.80 | 75.74 ± 26.11 | 80.27 ± 20.98 |
| Mini-Mental State Exam (MMSE) | 28.88 ± 1.52 | 28.82 ± 1.52 | 28.88 ± 1.51 | 28.70 ± 1.60 | **28.88 ± 1.51** | **28.26 ± 1.78** |
| Digit Symbol | **47.58 ± 12.53** | **45.86 ± 12.31** | 47.28 ± 12.51 | 45.75 ± 12.29 | **47.23 ± 12.54** | **43.94 ± 10.57** |
| Digit Span Forward | 8.34 ± 2.12 | 8.46 ± 2.22 | 8.38 ± 2.15 | 8.31 ± 2.15 | 8.39 ± 2.14 | 7.75 ± 2.25 |
| Digit Span Backward | 6.64 ± 2.28 | 6.64 ± 2.32 | 6.66 ± 2.29 | 6.47 ± 2.22 | 6.66 ± 2.29 | 6.06 ± 2.08 |

Notes: Values are expressed as mean ± standard deviation, or number (percentage%). Female sex is represented as total number of sample and percentage of sample. SCD- = cognitively normal older adults without subjective cognitive decline. SCD+ = cognitively older adults with subjective cognitive decline. CDR = clinical dementia rating. Bolded values represent significant differences in means between the SCD- and SCD+ groups.

**Table 2.** Demographic and variable characteristics for all SCD definitions for older adults in the MCI group.

| MCI | Patient |  | Informant |  | Clinician |  |
| --- | --- | --- | --- | --- | --- | --- |
|  | SCD- | SCD+ | SCD- | SCD+ | SCD- | SCD+ |
| Total number | 170 | 670 | 160 | 680 | 47 | 793 |
| Age | 70.33 ± 6.67 | 69.77 ± 6.45 | 68.91 ± 6.72 | 70.11 ± 6.42 | 70.06 ± 6.87 | 69.87 ± 6.47 |
| Female Sex | 80 (47%) | 300 (45%) | 92 (58%) | 288 (42%) | 30 (64%) | 350 (44%) |
| Education | 14.38 ± 4.15 | 14.51 ± 3.92 | 14.43 ± 4.29 | 14.50 ± 3.88 | 12.53 ± 5.37 | 14.60 ± 3.84 |
| CDR | 1.50 ± 1.23 | 1.56 ± 1.15 | **1.13 ± 1.07** | **1.65 ± 1.17** | **0.96 ± 0.67** | **1.58 ± 1.18** |
| Animal Fluency | 15.50 ± 5.46 | 15.54 ± 5.10 | 15.95 ± 5.31 | 15.44 ± 5.14 | 16.39 ± 4.65 | 15.48 ± 5.20 |
| Vegetable Fluency | 10.65 ± 3.94 | 10.59 ± 3.87 | **11.46 ± 4.16** | **10.41 ± 3.79** | 11.60 ± 3.70 | 10.54 ± 3.89 |
| Trail-Making A | 50.24 ± 25.94 | 45.87 ± 26.03 | 49.73 ± 27.47 | 46.08 ± 25.71 | 54.22 ± 29.78 | 46.29 ± 25.77 |
| Trail-Making B | 94.96 ± 29.32 | 91.54 ± 27.69 | 91.87 ± 27.30 | 92.16 ± 28.13 | 99.23 ± 25.95 | 91.77 ± 28.04 |
| Mini-Mental State Exam (MMSE) | 26.45 ± 4.00 | 26.98 ± 2.66 | 26.50 ± 4.20 | 26.95 ± 2.63 | 26.85 ± 2.92 | 26.87 ± 2.99 |
| Digit Symbol | 34.64 ± 14.73 | 36.64 ± 12.86 | 35.59 ± 14.44 | 36.39 ± 13.01 | 32.87 ± 14.42 | 36.46 ± 13.18 |
| Digit Span Forward | 7.14 ± 2.25 | 7.53 ± 2.39 | 7.06 ± 2.54 | 7.54 ± 2.32 | 6.73 ± 2.73 | 7.50 ± 2.34 |
| Digit Span Backward | 5.17 ± 2.09 | 5.64 ± 2.15 | 5.20 ± 2.20 | 5.62 ± 2.13 | 5.04 ± 2.03 | 5.57 ± 2.15 |

Notes: Values are expressed as mean ± standard deviation, or number (percentage%). Female sex is represented as the total number of sample and percentage of sample. SCD- = cognitively normal older adults without subjective cognitive decline. SCD+ = cognitively older adults with subjective cognitive decline. CDR = clinical dementia rating. Bolded values represent significant differences in means between the SCD- and SCD+ groups.

**Table 3.** Demographic and variable characteristics for all SCD definitions for older adults in the AD group.

| AD | Patient |  | Informant |  | Clinician |  |
| --- | --- | --- | --- | --- | --- | --- |
|  | SCD- | SCD+ | SCD- | SCD+ | SCD- | SCD+ |
| Total number | 280 | 1480 | 85 | 1675 | 16 | 1744 |
| Age | **71.69**<br>± **6.77** | **70.27**<br>± **7.19** | **67.33**<br>± **6.97** | **70.65**<br>± **7.12** | 70.69<br>± 6.59 | 70.49<br>± 7.15 |
| Female Sex | 157 (56%) | 829 (56%) | 52 (61%) | 934 (56%) | 7 (44%) | 979 (56%) |
| Education | **13.18**<br>± **4.65** | **14.72**<br>± **3.86** | 14.92<br>± 4.24 | 14.45<br>± 4.02 | 13.88<br>± 5.63 | 14.48<br>± 4.02 |
| CDR | **7.77**<br>± **5.63** | **4.83**<br>± **3.38** | **1.63**<br>± **2.03** | **5.48**<br>± **4.30** | **0.81**<br>± **0.87** | **5.34**<br>± **4.30** |
| Animal Fluency | **10.01**<br>± **6.46** | **11.82**<br>± **5.82** | **15.44**<br>± **6.57** | **11.35**<br>± **5.85** | **15.93**<br>± **3.53** | **11.51**<br>± **5.96** |
| Vegetable Fluency | **6.71**<br>± **4.85** | **7.89**<br>± **4.44** | **11.68**<br>± **5.15** | **7.51**<br>± **4.39** | **11.53**<br>± **3.80** | **7.68**<br>± **4.51** |
| Trail-Making A | **77.06**<br>± **46.65** | **64.10**<br>± **40.89** | 58.14<br>± 41.38 | 66.30<br>± 41.94 | 51.13<br>± 32.21 | 66.02<br>± 42.01 |
| Trail-Making B | **101.86**<br>± **28.41** | **94.17**<br>± **29.42** | **86.29**<br>± **30.59** | **95.67**<br>± **29.20** | 90.83<br>± 32.07 | 94.99<br>± 29.37 |
| Mini-Mental State Exam (MMSE) | **18.34**<br>± **8.08** | **21.54**<br>± **6.37** | **26.29**<br>± **3.73** | **20.79**<br>± **6.78** | **26.20**<br>± **3.45** | **21.0**<br>± **6.77** |
| Digit Symbol | **23.93**<br>± **16.25** | **28.86**<br>± **16.01** | **36.59**<br>± **17.65** | **27.74**<br>± **15.93** | 36.33<br>± 14.04 | 28.08<br>± 16.13 |
| Digit Span Forward | 6.26<br>± 2.50 | 6.96<br>± 2.40 | 7.25<br>± 1.83 | 6.84<br>± 2.12 | 7.47<br>± 1.99 | 6.85<br>± 2.43 |
| Digit Span Backward | 3.76<br>± 2.22 | 4.62<br>± 2.31 | 5.22<br>± 2.23 | 4.45<br>± 2.31 | 5.20<br>± 2.21 | 4.48<br>± 2.31 |

Notes: Values are expressed as mean ± standard deviation, or number (percentage%). Female sex is represented as the total number of sample and percentage of sample. SCD- = cognitively normal older adults without subjective cognitive decline. SCD+ = cognitively older adults with subjective cognitive decline. CDR = clinical dementia rating. Bolded values represent significant differences in means between the SCD- and SCD+ groups.

### 2.3 NACC Cognitive Variables

To evaluate the differences between patient endorsed, informant endorsed, and clinician endorsed SCD and their influence on cognitive performance, we systematically assessed multiple variables reflecting different domains of cognition. We used the CDRSUM variable to assess overall cognitive and functional ability, representing scores on the Clinical Dementia Rating - Sum of Boxes. The NACCMMSE variable was also used as a measure of global cognition, representing scores on the Mini-Mental State Exam (MMSE). Measures of language fluency included the variables ANIMALS and VEG, which represent the animal and vegetable fluency tests. Measures of executive function were derived from the TRAILA and TRAILB variables, representing the Trail-making tests A and B. The WAIS variable, representing the Wechsler Adult Intelligence Scale - Digit Symbol, was used as a measure of processing speed and executive functioning. Finally, the DIGIF and DIGIB variables, representing the forward and backward digit span tests, were used as measures of short-term memory. For the CDRSUM, TRAILA and TRAILB variables, higher scores indicate worse cognitive performance. For the ANIMALS, VEG, NACCMMSE, WAIS, DIGIF and DIGIB variables, lower scores indicate worse impairment.

### 2.4 Statistical Analyses

Analyses were performed using MATLAB R2024b. Demographic and cognitive characteristics were analysed using independent samples t-tests and analysis of variance. To investigate the relationship between SCD status and cognitive performance for each type of SCD, we performed a series of linear regression models, including sex, education, age as covariates. The main effect of interest was SCD_Status, reflecting the comparison between SCD+ and SCD-. Separate linear regressions were completed for the patient, informant, and clinician SCD definition for each cognitive outcome:

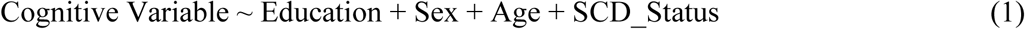

Additional longitudinal analyses were conducted to investigate the relationship between SCD status and cognitive performance over time. Linear mixed-effects analyses were conducted to investigate whether SCD status would influence future cognitive performance, including sex, education, age at baseline, and time from baseline as covariates. Participant ID was included as a categorical random effect. Separate analyses were completed for the three SCD definitions for each cognitive status and each cognitive outcome:

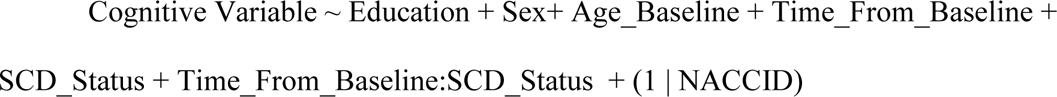

In these analyses, the main effect of interest was the Time_From_Baseline: SCD_Status interaction, indicating whether the slopes of cognitive decline were significantly different between SCD+ and SCD-groups. All p-values are reported as corrected values with significance determined by false discovery rate (FDR) controlling method to account for the multiple comparisons performed [33].

## 3. Results

### 3.1 Demographics

Fig. 1 displays a Venn diagram of the overlap between the different types of SCD for the NC group. Of the 1690 normal control participants, 552 (33%) participants were identified as SCD+ with at least one definition, whereas 1138 (67%) were classified as SCD-. Of the SCD+ participants, 118 (21%) were SCD+ across two definitions of SCD, with the majority of the overlap found between the patient and informant definitions (19%). Only 13 (0.02%) participants were SCD+ across all three definitions. As expected, the majority of the cases in the MCI and AD group were classified as SCD+.

**Figure 1:**
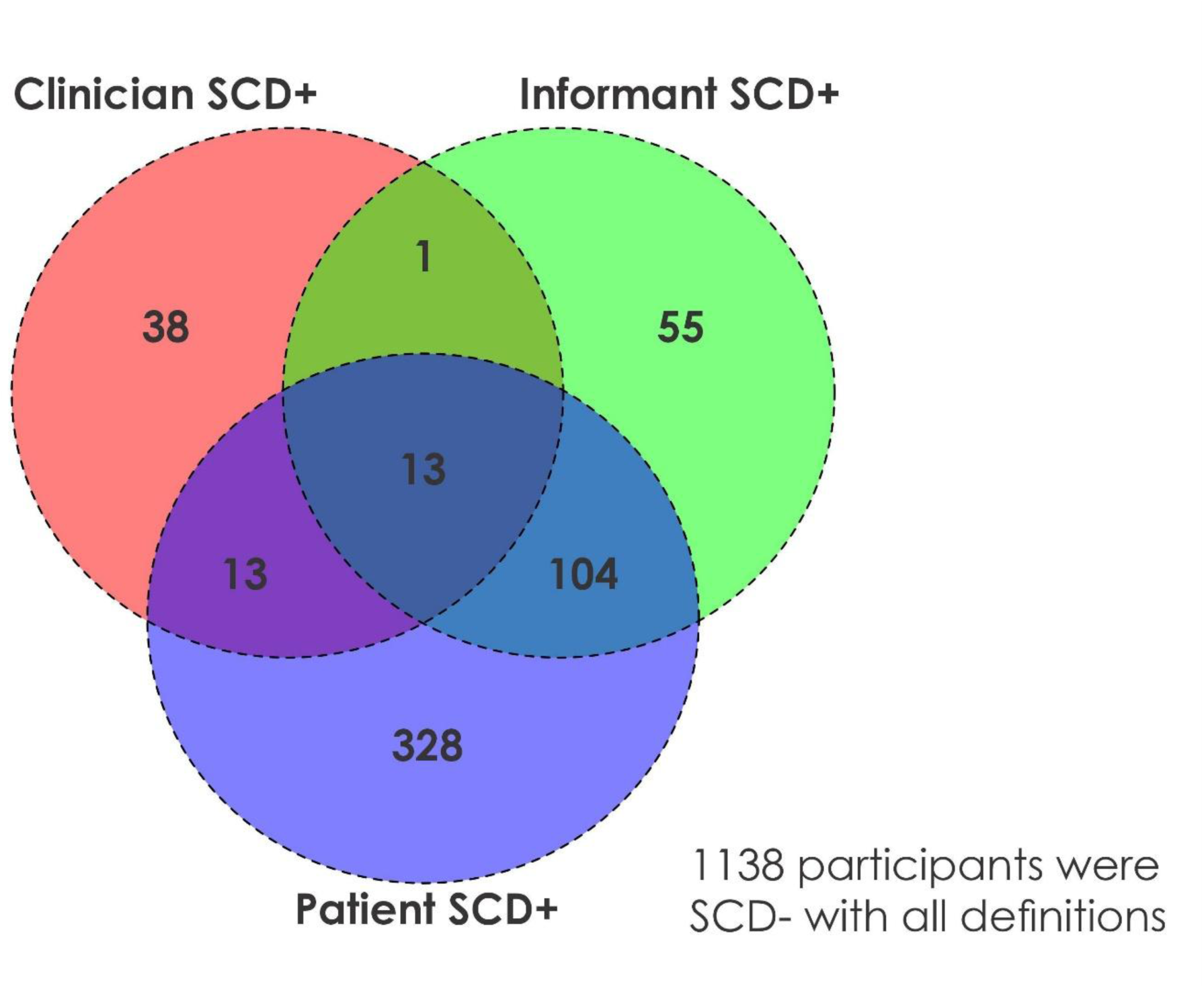
Venn Diagram representing the overlap of subjective cognitive decline (SCD) diagnosis between the three definitions of SCD in the normal control (NC) group. Notes: There was a total of 1690 participants in the sample, 1138/1690 (67%) were SCD-with all definitions. The remaining 552 participants are shown in the Venn Diagram with the number of participants. For the three SCD definitions, there were 458 SCD+ subjects defined by the patient definition, 173 SCD+ defined by the informant definition, and 65 SCD+ defined by the clinician definition of SCD. Finally, only 13 of the 552 SCD+ subjects are common between the three definitions.

Table 1 shows the demographic information and variable characteristics for NC participants using the three SCD definitions. No statistically significant differences in mean age or education between groups were observed between SCD+ and SCD-participants for all SCD definitions. No statistically significant differences in male to female ratio were observed for the patient and clinician definitions of SCD, but for informant SCD+, a higher proportion of males compared to females in the SCD+ group was present (*x*^2^ =11.0, *p* < .001).

Table 2 shows the demographic information and variable characteristics for MCI participants using the three SCD definitions. No statistically significant differences in mean age or education between groups were observed between SCD+ and SCD-participants for all SCD definitions. No statistically significant difference in male to female ratio was observed for the patient definition of SCD. However, there was a significant difference in male to female ratio for the informant (*x*^2^ = 12.0, *p* < .001) and clinician (*x*^2^ = 6.95, *p* = .008) definitions of SCD, with a higher proportion of males compared to females in the SCD+ group for both definitions.

Table 3 shows the demographic information for AD participants using the three SCD definitions. No significant differences in age were observed for the clinician definition; however, for informant SCD, those with SCD+ were older than SCD- (*t* = 4.20, *p* < .001). Conversely, for the patient definition, participants were older in the SCD-group than SCD+ (*t* = -3.06, *p* = .002). No significant differences in education level were observed for the informant and clinician definitions, but participants in the patient SCD+ group were more highly educated than those in the SCD-group (*t* = 5.98, *p <* .001). No statistically significant differences in male to female ratio were observed between SCD+ and SCD-participants for all definitions.

### 3.2 Cross-sectional analysis

No significant differences were found between the cognitive profiles of the self-reported SCD- and SCD+ in the NC group at baseline. Compared to MCI participants with no self-reported complaints, MCI participants with self-reported SCD displayed worse performance on the Vegetable Fluency (*t* = -2.30, *p* = 0.02) measure. Compared to AD participants with no complaints, AD participants with self-reported SCD showed worse performance on the CDR-SB (*t* = 3.13, *p* = .002), Animal (*t* = -2.77, *p* = .005) and Vegetable Fluency (*t* = -2.88, *p* = .004), and MMSE (*t* = -2.63, *p* = .008) measures. All results for self-reported SCD in the MCI and AD groups remained statistically significant after applying an FDR correction.

Compared to NC participants with no informant complaints, NC participants with informant endorsed SCD showed worse baseline performance on the Animal Fluency (*t* = -2.27, *p* = .023) and WAIS (*t* = -2.07, *p* = .040). Compared to MCI participants with no complaints, MCI participants with informant endorsed SCD showed worse performance on the CDR (*t* = 2.15, *p* = .032) and Vegetable Fluency (*t* = -2.46, *p* = 0.01). Compared to AD participants with no complaints, AD participants with informant endorsed SCD showed worse performance on the CDR (*t* = 3.30, *p* <.001), Animal (*t* = -2.90, *p* = .003) and Vegetable (*t* = -3.10, *p* = .002) Fluency, MMSE (*t* = -2.82, *p* = .005), and WAIS (*t* = -2.09, *p* = .037). All results for informant SCD in all groups survived FDR correction.

Compared to NC participants with no positive clinician SCD report, NC participants with clinician SCD showed lower performance on the Animal Fluency (*t* = -2.93, *p* = .003), Trail-making A (*t* = 8.98, *p* < .001), MMSE (*t* = -3.77, *p* < .001), and Forward (*t* = -2.44, *p* = .015) and Backward (*t* = -2.35, *p* = .019) Digit Span. Compared to MCI participants with no complaints, MCI participants with clinician SCD showed lower performance on the Vegetable Fluency (*t* = - 2.34, *p* = 0.02) task. Compared to AD participants with no complaints, AD participants with clinician SCD showed worse outcomes on the CDR (*t* = 3.22, *p* = .001), Animal (*t* = -2.80, *p* = .005) and Vegetable (*t* = -2.95, *p* = .003) Fluency, MMSE (*t* = -2.73, *p* = .006), and WAIS (*t* = - 2.02, *p* = .044). All results for informant SCD in all groups survived FDR correction.

### 3.3 Longitudinal analyses

Table 4 summarizes the results of the linear mixed-effects analysis. Figures 2 & 3 show the predicted longitudinal CDR-SB trajectory for SCD+ and SCD-in the NC group, and in the MCI group, respectively. In the NC group, we found that having a positive self-report of SCD was related to increased cognitive decline over time on the CDR-SB (*t* = 3.19, *p* = .001), Animal (*t* = -2.53, *p* = .011) and Vegetable (*t* = -2.94, *p* = .003) Fluency, MMSE (*t* = -2.78, *p* = .005), Forward (*t* = -6.23, *p* < .001) and Backward (*t* = -6.50, *p* < .001) Digit Span compared to the SCD-group. Conversely, we found that a positive self report of SCD was related to better performance over time on the Trail-Making A (*t* = -2.70, *p* = .007) and B (*t* = -2.34, *p* = .019) tests compared to the SCD-group. In the MCI group, a positive self-report of SCD was related to increased cognitive decline over time on the CDR-SB (*t* = 3.04, *p* = .002) and Animal Fluency (*t* = -2.19, *p* = .028) compared to the SCD-control group. In the AD group, a positive self-report of SCD was related to increased decline over time on the CDR-SB (*t* = 2.95, *p* = .003) compared to the SCD-group. All results for self-reportSCD in all groups remained statistically significant after applying FDR correction for multiple comparisons.

**Table 4:**
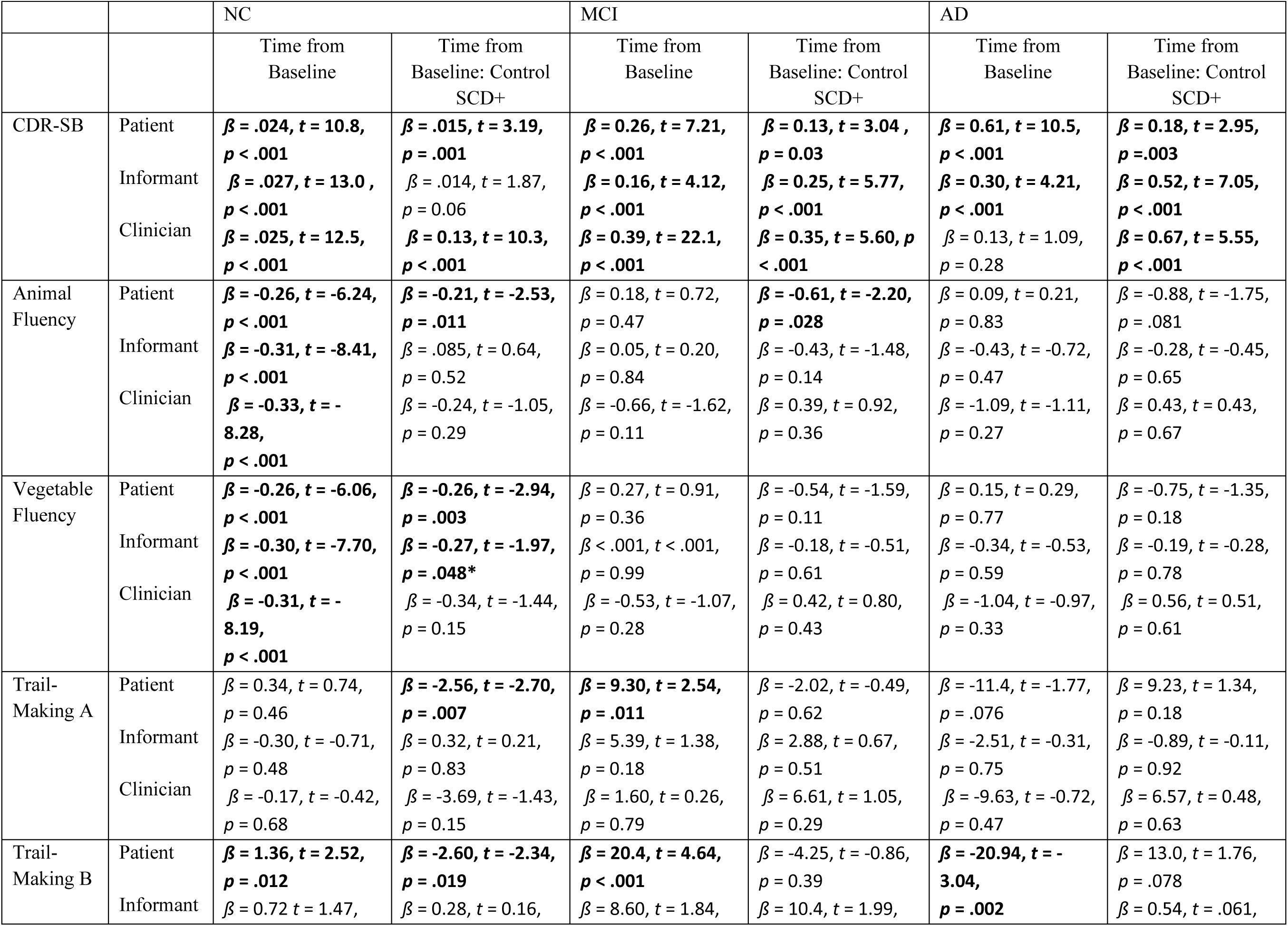
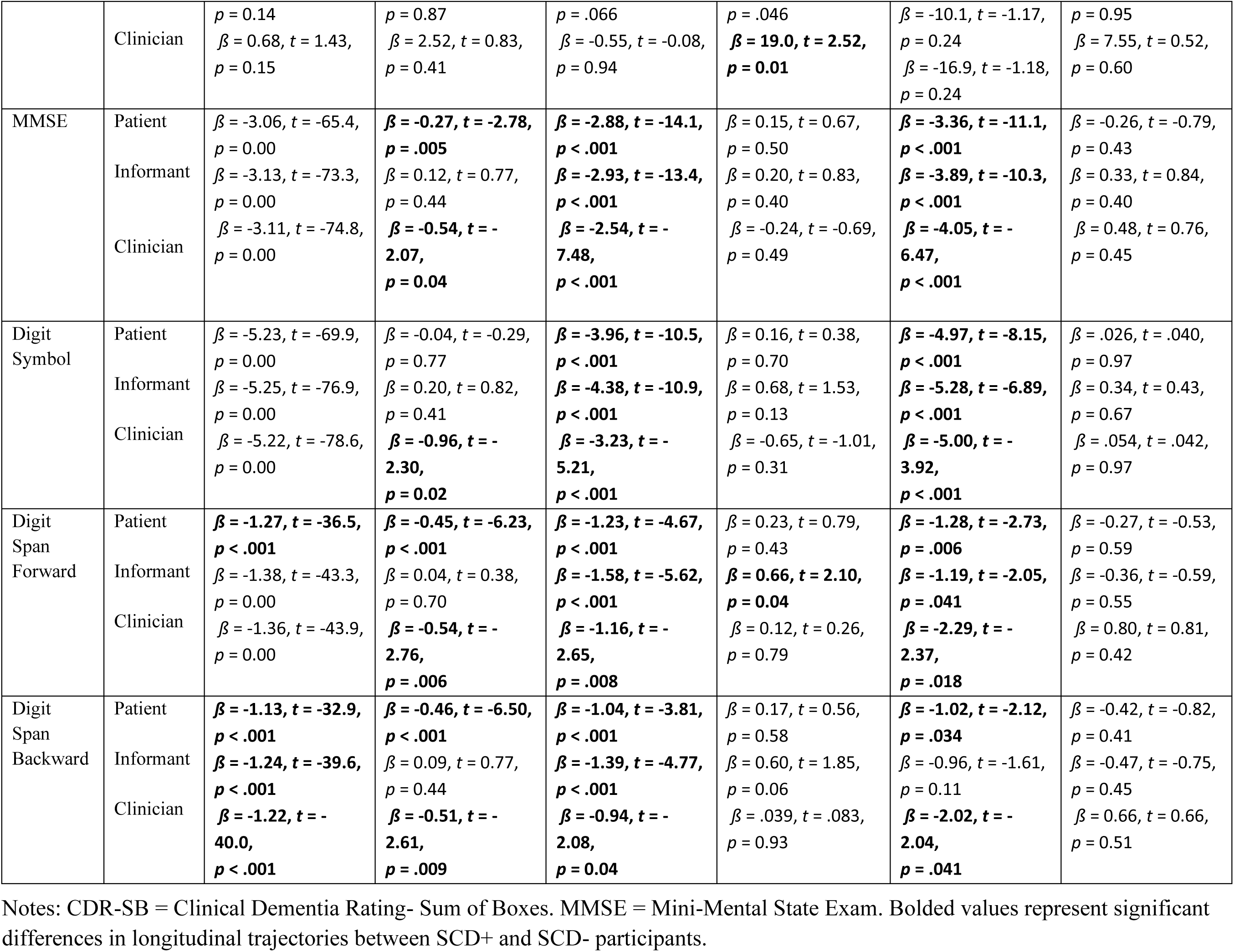
Linear mixed effects model results showing longitudinal trajectories in cognitive performance between SCD+ and SCD-participants for the three definitions of SCD.

**Figure 2:**
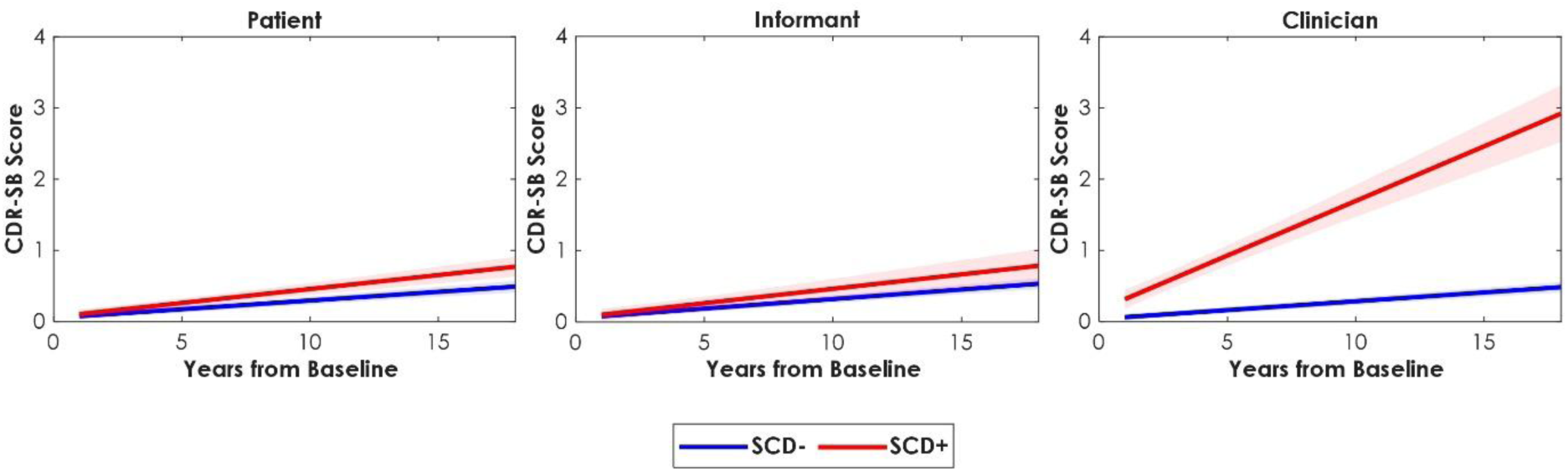
Predicted longitudinal CDR-SB trajectory for SCD+ and SCD-in the normal control group. Notes: CDR-SB = Clinical Dementia Rating - Sum of Boxes. SCD-= Older adult without subjective cognitive decline. SCD+ = Older adult with subjective cognitive decline. The patient and clinician definitions of SCD showed a significant difference in cognitive trajectories between SCD+ and SCD-, with SCD+ displaying worse cognitive trajectories.

**Figure 3:**
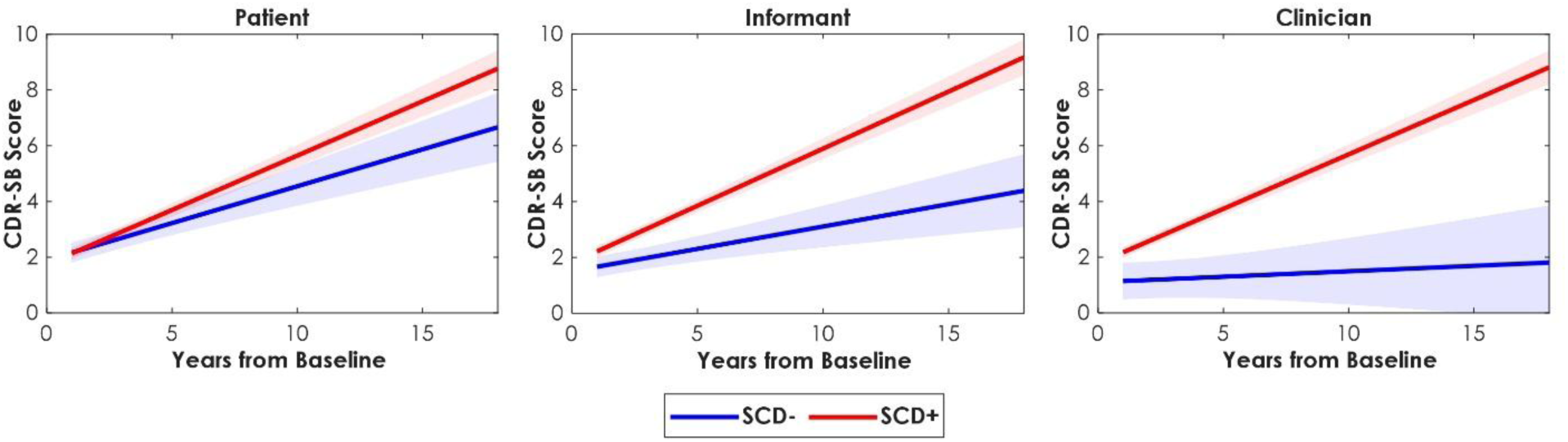
Predicted longitudinal CDR-SB trajectory for SCD+ and SCD-in the mild cognitive impairment group. Notes: CDR-SB = Clinical Dementia Rating - Sum of Boxes. SCD-= Older adult without subjective cognitive decline. SCD+ = Older adult with subjective cognitive decline. All three SCD definitions showed a significant difference in cognitive trajectories between SCD+ and SCD-, with SCD+ displaying worse cognitive trajectories.

In the NC group, we found that a positive informant SCD report was related to increased cognitive decline over time on the Vegetable Fluency (*t* = -1.97, *p* = .048) compared to the SCD-informant control group, although this result did not survive FDR correction. In the MCI group, a positive report of informant SCD was related to increased cognitive decline over time on the CDR-SB (*t* = 5.77, *p* < .001), Trail-Making B (*t* = 1.99, *p* = .046), and the Forward Digit Span (*t* = 2.10, *p* = .035) compared to the SCD-group. In the AD group, a positive report of informant SCD was related to increased cognitive decline over time on the CDR-SB (*t* = 7.05, *p* < .001) compared to the SCD-group. All results for informant SCD in the MCI and AD groups remained statistically significant after applying an FDR correction for multiple comparisons.

In the NC group, we found that a positive report of clinician SCD was related to increased cognitive decline over time on the CDR-SB (*t* = 10.3, *p* < .001), MMSE (*t* = -2.07, *p* = .038), WAIS (*t* = -2.30, *p* = .022), Forward (*t* = -2.76, *p* =.006) and Backward (*t* = -2.61, *p* = .009) Digit Span compared to the SCD-group. In the MCI group, we found that a positive clinician report of SCD was related to increased cognitive decline over time on the CDR-SB (*t* = 5.60, *p* = < .001) and Trail-Making B (*t* = 2.52, *p* = .012) compared to the SCD-group. In the AD group, we found that a positive clinician report of SCD was related to increased cognitive decline over time on the CDR-SB (*t* = 5.55, *p* = < .001) compared to the SCD-group. All results for clinician SCD in all groups remained statistically significant after the FDR correction.

## Discussion

Despite SCD’s relationship to cognitive decline [2,3,34], previous literature lacks a definitive understanding of the role of different definitions of SCD, including patient, informant, and clinician, as indicators of cognitive decline. The current study investigated differences in cognition between SCD+ and SCD-participants across three definitions of SCD and three diagnostic groups, to explore the differences between each SCD definition at baseline and over time. We found notable differences in SCD endorsement across groups. Importantly, while baseline patient self-report of SCD was not associated with cognition in any domain in the NC group, it was associated with significantly increased rates of cognitive decline. In NC, the informant definition of SCD was associated with worse cognitive performance at baseline, but the effects did not persist longitudinally. The clinician definition of SCD was associated with worse cognition at baseline and longitudinal cognitive decline in NC. In MCI and AD, all three SCD definitions were associated with worse cognition at baseline and longitudinal cognitive decline.

In NC, the patient SCD definition was not associated with worse cognition at baseline, but was associated with worse cognitive decline in language, global cognition, short-term memory, and overall functional ability. Our longitudinal results are supported by previous research [15], which found that SCD+ cognitively healthy adults exhibited greater decline across global cognition and episodic memory. They also found that while some cognitive domains did not show differences between SCD+ and SCD-at baseline, they showed differences longitudinally [15]. These results suggest that self-reported SCD is an important indicator for cognitive decline and may be more strongly associated with change over time than baseline cognition [3]. In MCI, the patient SCD definition was associated with worse cognition in language at baseline, and with longitudinal decline in language and overall cognitive/functional ability. These results are inconsistent with a previous study which observed that self-reported decline was not related to worse cognitive performance at baseline for MCI in language or overall cognitive/functional ability [32]. These differences may be related to sample size differences between the studies (with ours having a larger sample size), and the differences in the way SCD was defined across studies (i.e., with a single question vs. with the Everyday Cognition Scale (ECog)). Interestingly, another study found that participants with self-reported SCD that were NC at baseline and progressed to MCI over 5 years showed lower scores on language measures on baseline neuropsychological tests [35]. Therefore, difficulties in language may start to emerge between the NC and MCI stage, and performance in language may be an important indicator of which individuals with SCD will experience cognitive decline. This finding is relevant, as semantic decline has been significantly associated with the onset of specific types of dementia, such as fronto-temporal, compared to other types [36]. Therefore, it is crucial to identify which type(s) of dementia one may be more at risk for to enable more effective management [2,36].

Informant SCD reports were associated with worse baseline cognition and longitudinal cognitive decline across various cognitive domains in both the NC and MCI group. In NC, informant reports were associated with worse cognition at baseline in language, executive function, and processing speed. These results are consistent with previous research which has suggested that informant SCD in NC is related to lower cognition in executive function [16,27], language [21], and processing speed [21] and longitudinal cognitive decline in executive functions and memory [17]. Although we observed relationships between baseline cognition and informant reports in NCs, there were no associations over time. This finding is also consistent with some evidence indicating that informant-reports may be more useful in patients with evidence of decline [2]. Together, these conflicting reports indicate that in NCs, informant reports reflect immediate cognitive function but do not predict change over time. Therefore, considering that patient SCD was associated with future decline, our results suggest that self reports are superior to informant reports when exploring longitudinal cognitive decline in cognitively healthy older adults.

In MCI, the informant definition of SCD was associated with worse cognition at baseline in language and overall functional ability, as well as longitudinal cognitive decline in executive function, short-term memory, and overall functional ability. These results are supported by previous research in MCI which showed that informant SCD was associated with cognitive performance [32]. Additionally, our results support the previous literature which suggests the usefulness of informant SCD in MCI, particularly because patient’s insight into their own cognition become less reliable as cognitive decline progresses [30,31,37]. Overall, our results highlight the importance of informant reports as an additional measure of cognitive concern, particularly in those with MCI.

Clinician SCD reports were associated with worse baseline cognition and longitudinal cognitive decline across various cognitive domains in both NC and MCI. In NC, the clinician SCD definition was associated with worse cognition at baseline in language, executive function, global cognition, and short-term memory. As well, it was associated with worse longitudinal cognitive decline in executive function, short-term memory, global cognition, and in overall functional ability. In MCI, the clinician SCD definition was associated with worse cognition in language at baseline, and with worse longitudinal cognitive decline in executive function and overall functional ability. To our knowledge, this study is the first to report on the relationship between clinician SCD and objective cognition at baseline and longitudinally. Therefore, these results show promising potential for the use of clinician SCD as a relevant early indicator for the risk of further cognitive decline in SCD+ individuals. Additionally, clinician SCD was associated with worse decline in many cognitive domains, which may be explained by the question used to define SCD, which asked about the patient’s overall cognition, providing some explanation for the number of significant associations across multiple domains. Future research should investigate the validity of clinician SCD as a predictor of cognitive decline in both cognitively healthy older adults and those with MCI, and continue to characterize clinician definitions of SCD through measures that assess overall cognition.

In AD, we observed that all three definitions of SCD were related to worse cognition at baseline and longitudinally. At baseline, we found that all three definitions of SCD were associated with worse cognition in global cognition and language. Additionally, we found that the informant and clinician definitions were associated with worse cognition in executive function and processing speed. The significant results in informant and clinician SCD support the idea that positive reports of cognitive complaints from an outside source are better indicators of objective decline as patient’s insight into their own cognition decreases [30,31,37], which is likely in AD [38]. The significant differences between definitions in AD could be explained by the fact that patient and informant SCD are related to faster rates of objective decline in multiple cognitive domains [17,21,27], and this pattern may remain consistent as decline progresses to later disease stages, leading to earlier cognitive decline in AD with SCD compared to AD without SCD. Our longitudinal results suggest that AD patients who have SCD using any definition of SCD are more likely to experience decline on a measure that evaluates everyday functional and cognitive performance [39], leading to worse decline in daily functioning compared to AD patients without SCD. The lack of longitudinal results in measures that target specific domains of cognition may be explained by the fact that AD patients experience decline in multiple cognitive domains [40], possibly leading to a convergence in the severity of symptoms between SCD+ and SCD-patients as the disease progresses. Our results in the AD group suggest additional support for SCD (particularly informant and clinician reports) as a good early indicator of cognitive decline.

While clinician SCD has been defined in the past [26], to our knowledge, this was the first study to consider the importance of clinician SCD reports when examining longitudinal cognitive change. However, there are a few limitations to the current study. First, patient and informant SCD were determined using only a memory-concern question, which does not account for overall cognition [3]. Therefore, future research should quantify SCD using comprehensive scales that assess concerns in a variety of domains such as the Everyday Cognition Scale [41] or the Cognitive Change Index [42] . Finally, all of our participants’ education levels were relatively high, potentially limiting the generalizability of our results to representative populations.

The current study examined the relationship between SCD and cognitive decline using three SCD definitions. These findings have significant implications for future research by revealing the pertinent role of clinician reports in predicting cognitive decline, and suggesting its use alongside patient and informant reports. In patients reporting SCD, the combination of the three SCD definitions would provide a comprehensive overview of the likelihood of cognitive decline, and possible conversion to MCI or dementia. Across all the cognitive domains observed, language was most commonly associated with worse cognition and cognitive decline across all definitions. Thus, we suggest that future research focus on monitoring declines in language in individuals with any definition of SCD.

## Data Availability

All data produced are available online at https://www.naccdata.org/

## Acknowledgments

The NACC database is funded by NIA/NIH Grant U24 AG072122. NACC data are contributed by the NIAfunded ADRCs: P30 AG062429 (PI James Brewer, MD, PhD), P30 AG066468 (PI Oscar Lopez, MD), P30 AG062421 (PI Bradley Hyman, MD, PhD), P30 AG066509 (PI Thomas Grabowski, MD), P30 AG066514 (PI Mary Sano, PhD), P30 AG066530 (PI Helena Chui, MD), P30 AG066507 (PI Marilyn Albert, PhD), P30 AG066444 (PI David Holtzman, MD), P30 AG066518 (PI Lisa Silbert, MD, MCR), P30 AG066512 (PI Thomas Wisniewski, MD), P30 AG066462 (PI Scott Small, MD), P30 AG072979 (PI David Wolk, MD), P30 AG072972 (PI Charles DeCarli, MD), P30 AG072976 (PI Andrew Saykin, PsyD), P30 AG072975 (PI Julie A. Schneider, MD, MS), P30 AG072978 (PI Ann McKee, MD), P30 AG072977 (PI Robert Vassar, PhD), P30 AG066519 (PI Frank LaFerla, PhD), P30 AG062677 (PI Ronald Petersen, MD, PhD), P30 AG079280 (PI Jessica Langbaum, PhD), P30 AG062422 (PI Gil Rabinovici, MD), P30 AG066511 (PI Allan Levey, MD, PhD), P30 AG072946 (PI Linda Van Eldik, PhD), P30 AG062715 (PI Sanjay Asthana, MD, FRCP), P30 AG072973 (PI Russell Swerdlow, MD), P30 AG066506 (PI Glenn Smith, PhD, ABPP), P30 AG066508 (PI Stephen Strittmatter, MD, PhD), P30 AG066515 (PI Victor Henderson, MD, MS), P30 AG072947 (PI Suzanne Craft, PhD), P30 AG072931 (PI Henry Paulson, MD, PhD), P30 AG066546 (PI Sudha Seshadri, MD), P30 AG086401 (PI Erik Roberson, MD, PhD), P30 AG086404 (PI Gary Rosenberg, MD), P20 AG068082 (PI Angela Jefferson, PhD), P30 AG072958 (PI Heather Whitson, MD), P30 AG072959 (PI James Leverenz, MD).

## Consent Statement

Written informed consent was obtained from participants or their study partner.

## Availability of data and materials

The data utilized in this study were also sourced from the National Alzheimer’s Coordinating Center (NACC) database (https://naccdata.org/), specifically drawing from the NACC Uniform Data Set (UDS) and MRI Data Set (Beekly et al., 2004; Besser, Kukull, Knopman, et al., 2018; Besser, Kukull, Teylan, et al., 2018).

